# Foundation Models for Quantitative Biomarker Discovery in Cancer Imaging

**DOI:** 10.1101/2023.09.04.23294952

**Authors:** Suraj Pai, Dennis Bontempi, Ibrahim Hadzic, Vasco Prudente, Mateo Sokač, Tafadzwa L. Chaunzwa, Simon Bernatz, Ahmed Hosny, Raymond H Mak, Nicolai J Birkbak, Hugo JWL Aerts

## Abstract

Foundation models represent a recent paradigm shift in deep learning, where a single large-scale model trained on vast amounts of data can serve as the foundation for various downstream tasks. Foundation models are generally trained using self-supervised learning and excel in reducing the demand for training samples in downstream applications. This is especially important in medicine, where large labeled datasets are often scarce. Here, we developed a foundation model for imaging biomarker discovery by training a convolutional encoder through self-supervised learning using a comprehensive dataset of 11,467 radiographic lesions. The foundation model was evaluated in distinct and clinically relevant applications of imaging-based biomarkers. We found that they facilitated better and more efficient learning of imaging biomarkers and yielded task-specific models that significantly outperformed their conventional supervised counterparts on downstream tasks. The performance gain was most prominent when training dataset sizes were very limited. Furthermore, foundation models were more stable to input and inter-reader variations and showed stronger associations with underlying biology. Our results demonstrate the tremendous potential of foundation models in discovering novel imaging biomarkers that may extend to other clinical use cases and can accelerate the widespread translation of imaging biomarkers into clinical settings.

## INTRODUCTION

Foundation models present a paradigm shift in deep learning wherein a model trained on vast amounts of unannotated data can serve as the foundation of a wide range of downstream tasks. Recently foundation models have provided unprecedented performance gains in language, vision, and several other domains^1^. In the field of natural language processing (NLP), for example, foundation models drive the successes of applications such as ChatGPT^2^, BERT^3^, and CLIP^4^. Similarly, foundation models, such as SimCLR^5^ and DINO^6^, have reported considerable success in computer vision applications.

Medicine represents a vast potential for foundation models as labeled data are scarce, while multimodal data, such as medical images, biologic, and clinical notes, are frequently collected in routine clinical care^7^. Indeed, different applications of foundation models, such as augmented surgical procedures, bedside decision support, interactive radiology reports, and note-taking, have been reported^8^.

While many studies investigating imaging-based biomarkers incorporate supervised deep learning algorithms into their models^9–11^, they are typically applied in scenarios where large datasets are available for training and testing. The quantity and quality of annotated data are strongly linked to the robustness of deep learning models. Access to large amounts of annotated data for specialized applications is often challenging and demands expertise, time, and labor. In such scenarios, many investigators fall back on traditional handcrafted or engineered approaches based on defined mathematical and statistical algorithms that analyze attributes like the shape and texture of objects in images, which limit the scope of discovery. This caveat is commonplace in many scenarios where insights from imaging-based biomarkers have great potential in informing clinical care.

Foundation models are generally pre-trained using self-supervised learning (SSL), a set of methods that leverage innate information available within data by learning generalized, task-agnostic representations (features) from large amounts of unannotated samples. Existing literature^12^ has suggested several strategies to pre-train networks to learn these representations. Approaches such as defining pre-text tasks that distort an image and attempt to reconstruct the original or contrastively learning similar representations for augmented views of the same image have primarily been investigated. Following pre-training, foundation models can be applied to task-specific problems, improving generalization, especially in tasks with small datasets. The expanding literature on SSL in medical imaging^13^ focuses primarily on two-dimensional images (X-ray, whole slide images, dermatology images, fundus images, etc.) and diagnostic applications. There is still limited evidence investigating whether SSL can help train foundation models that learn general, robust, and transferrable representations that can act as imaging biomarkers, especially prognostic, for tasks of clinical relevance.

In this study, we investigated whether foundation models pre-trained using self-supervised learning can improve the development of deep learning-based imaging biomarkers. The foundation model was pre-trained on 11,467 diverse and annotated lesions identified on computed tomography (CT) imaging from 2,312 unique patients^14^. The model was first technically validated on the classification of anatomical site lesions (use-case 1). Subsequently, it was applied to two distinct clinically relevant applications: the development of a diagnostic biomarker that predicts the malignancy of lung nodules (use-case 2) and a prognostic biomarker for non-small cell lung cancer tumors in confirmed cancer cases (use-case 3). We evaluated two distinct approaches of how a pre-trained foundation model can be incorporated into training pipelines for downstream tasks, a direct approach of using the foundation model as a feature extractor combined with a linear classifier and another approach where the foundation model is fine-tuned through deep learning. The performance of the foundation model approaches was then evaluated and compared to conventional supervised approaches in the three clinical use cases. Our analysis delves into limited data scenarios, evaluating test-retest and inter-reader stability, determining explainability and interpretability through deep-learning attribution methods, and exploring biological associations with gene expression data. Our results demonstrate the potential of foundation models in discovering novel imaging biomarkers and their particular strength in applications with limited datasets. This evidence may extend to other clinical use cases and imaging modalities and can accelerate the widespread development and translation of imaging biomarkers into clinical settings. To allow effortless incorporation, external evaluation, and validation, we are providing open access to the foundation model along with reproducible workflows.

## RESULTS

We developed a foundation deep learning model using SSL and tested the model’s performance in three distinct use cases. The study design and the pre-training process are outlined in **Fig. 1**. We developed the foundation model using a dataset with 11,467 annotated CT lesions identified from 2,312 unique patients. Lesion findings were diverse and included multiple lesions, such as lung nodules, cysts, and breast lesions, among numerous others. A task-agnostic contrastive learning strategy was used to pre-train the model on these lesion findings (see **Fig. 1a**), which subsequently was evaluated in three diverse clinical applications and five distinct datasets (see **Fig. 1b**).

**Figure 1.**
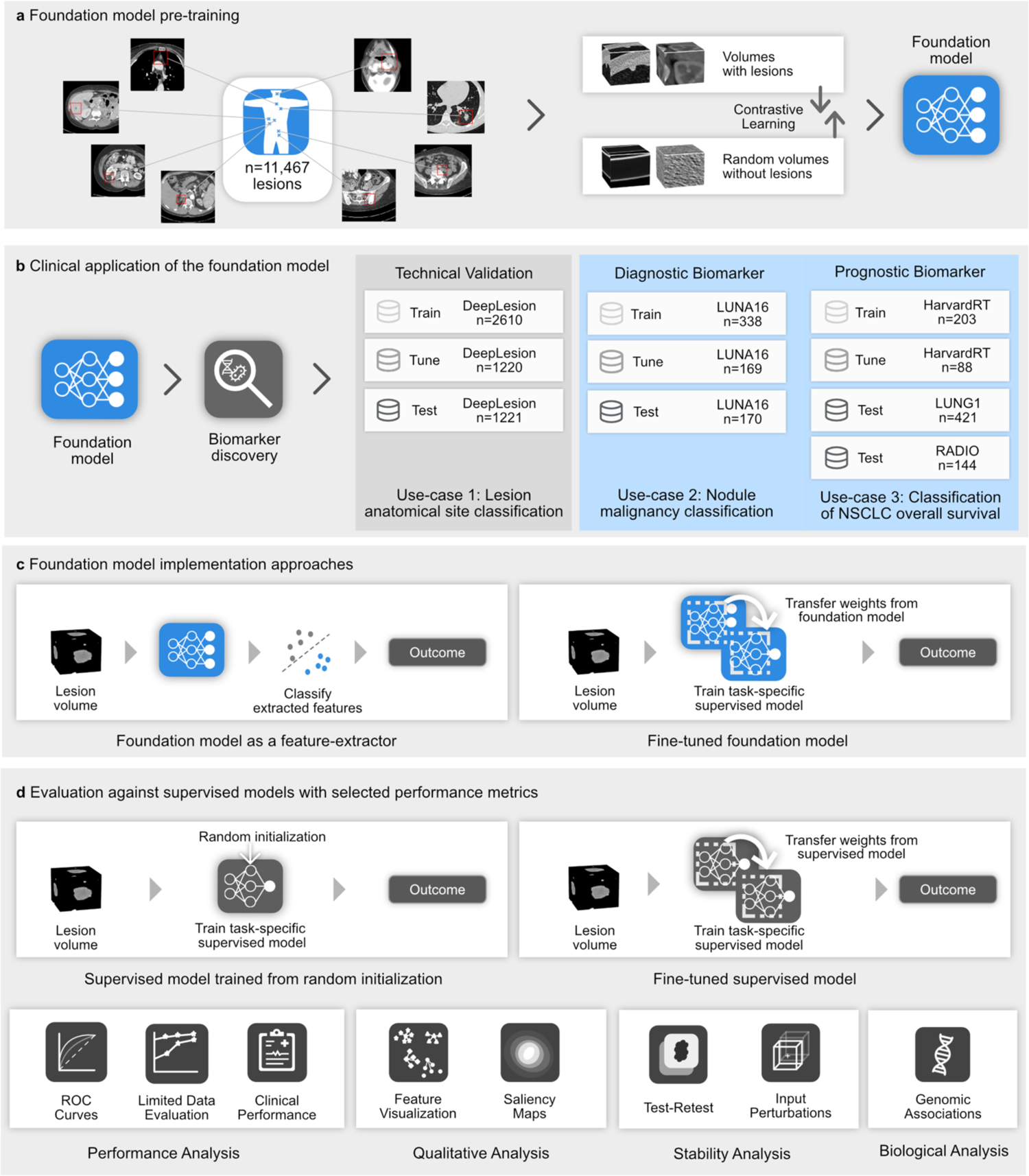
General overview of the study. a. Foundation model pre-training. **A** foundation model, specifically a deep convolutional encoder model, was pre-trained by contrasting volumes with and without lesions. **b. Clinical application of the foundation model.** The foundation model was used to extract biomarkers and then evaluated for three classification tasks on diverse datasets. **c. Foundation model implementation approaches** The foundation model was adapted to specific use cases by extracting features or through fine-tuning (left). **d. Evaluation against supervised models with selected performance metrics.** We compared the performance of the foundation models against conventional supervised implementations, trained from random intialization (left) and fine-tuned from a different task (right). The comparison was made through several criteria for the different use cases, including quantitative performance, stability, and biological analysis. Biological, clinical, and stability analyses are limited to use case 2 due to the availability of associated data.

### Lesion anatomical site classification (Use-case 1)

As a technical validation of the performance of the foundation model, we selected an in-distribution task (i.e., sourced from the same cohort as that of the foundation model pre-training) on 5,051 annotated lesions (see Use-case 1 in **Fig. 1b**). These specific lesions, however, were not included in the pre-training data. Classification models were developed to predict the correct anatomical site using a training and tuning dataset totaling 3,830 lesions. On an independent test set of 1,221 lesions, we evaluated the performance of two different implementations of the foundation model (see **Fig. 1c**).

We found that the foundation model approaches significantly outperformed the current standard supervised approach using a randomly initialized model (i.e., random initialization of weights; see **Fig. 1d**) in terms of balanced accuracy (BA) and mean average precision (mAP) (see **Fig. 2a, b**). When comparing classification performances, the foundation features-based classifier (0.779 [95% CI 0.749-0.809], p<0.01) and the fine-tuned foundation model (0.804 [95% CI 0.773-0.834], p<0.01), significantly improved BA (p<0.01) over the supervised model (0.72, [95% CI 0.689-0.750], p<0.01) (see **Fig. 2a**). In terms of mAP, the fine-tuned foundation model (0.856, [95% CI 0.828-0.886], p<0.01) provided a significant (p<0.01) performance benefit over the supervised model (mAP=0.818 [95% CI 0.779-0.847], p<0.01) (see **Fig. 2b**)

**Figure 2.**
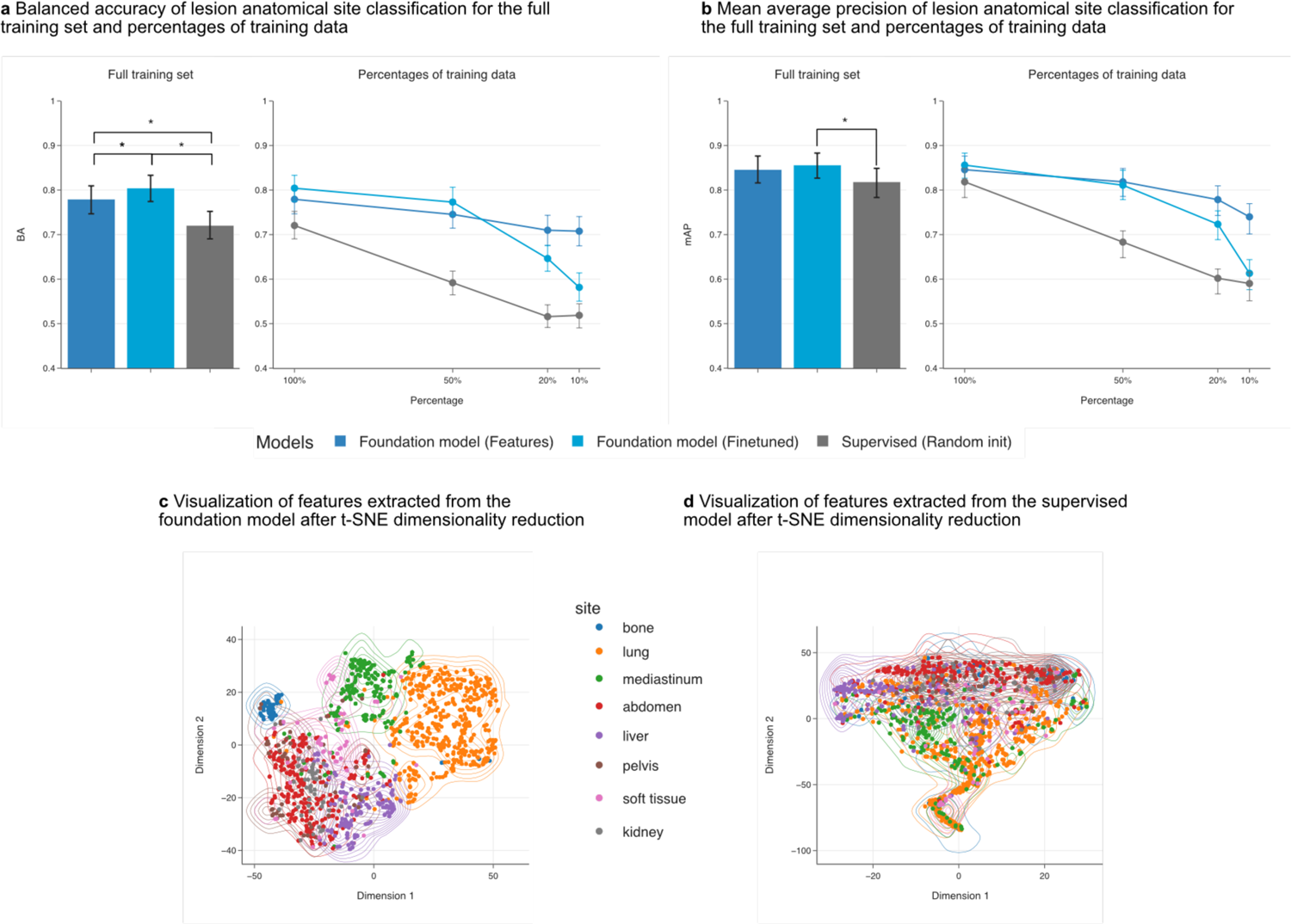
Performance of foundation model for lesion anatomical site classification. We compared foundation model adaptation approaches against a supervised model using balanced accuracy (**a**) and mean average precision (**b**). We show performance on these metrics computed across the eight anatomical sites for the full training set and when the training data percentage is decreased to 50%, 20%, and 10%. Error bars in (**a**) and (**b**) show 95% confidence intervals of the estimates. Visual representation of the features generated from the independent test-set for identifying lesion anatomical sites, using **c** the foundation model as a feature extractor, and **d** the supervised model. For (**c)** and (**d**), the x-axis corresponds to dimension 1, and the y-axis to dimension 2 of the t-SNE dimensionality reduction. The density contours belonging to each class are underlaid for (**c**) and (**d**) to highlight separability between classes in the feature space.

The performance advantage of the foundation model was even stronger in limited data scenarios (see **Fig. 2a, b**). When we reduced training data to 50% (n=2526), 20% (n=1010), and 10% (n=505), the foundation model as a feature extractor significantly improved BA and mAP over the supervised model. The fine-tuned foundation model also significantly improved over the supervised model but failed to improve when training data was reduced to 10%. Individual comparisons between each model at different data percentages can be found in the supplementary material (see **Extended Data Table 1**).

To investigate feature separability, which indicates how well features can discriminate between anatomical sites, we used dimensionality reduction methods to visualize features generated on the test set by the foundation and the trained supervised models. The features from the foundation model produced semantically separable clusters for each anatomical site, while features from the supervised model showed poor separability (see **Fig. 2c-d**). Of note, unlike the supervised model, the foundation model was not exposed to anatomical site information during training.

### Nodule malignancy prediction (Use case 2)

To assess the robustness of the foundation model, we chose an out-of-distribution task (i.e., belonging to a different cohort than that of the foundation model training data) involving predicting the malignancy of lung nodules from the LUNA16 dataset (see Use-case II in **Fig. 1b**). We conducted our training on a labeled subset of 507 lung nodules with indications of malignancy suspicion. On an independent test set of 170 nodules, we evaluated the performance of the two foundation model implementations and two supervised learning approaches - random initialization and fine-tuning from another supervised model. The model trained in use case 1 was chosen for the supervised fine-tuning.

The approach of fine-tuning the foundation model resulted in significant (p<0.01) superiority over both the supervised learning approaches (see **Fig. 3a, b**). The fine-tuned foundation model achieved an area-under receiver operating curve (AUC) of 0.944 (95% CI 0.914-0.982, p<0.01) and mAP of 0.952 (95% CI 0.926-0.986, p<0.01) compared to the fine-tuned supervised model’s AUC of 0.857 (95% CI 0.806-0.918, p<0.01) and mAP of 0.874 (95% CI 0.822-0.936, p<0.01).

**Figure 3.**
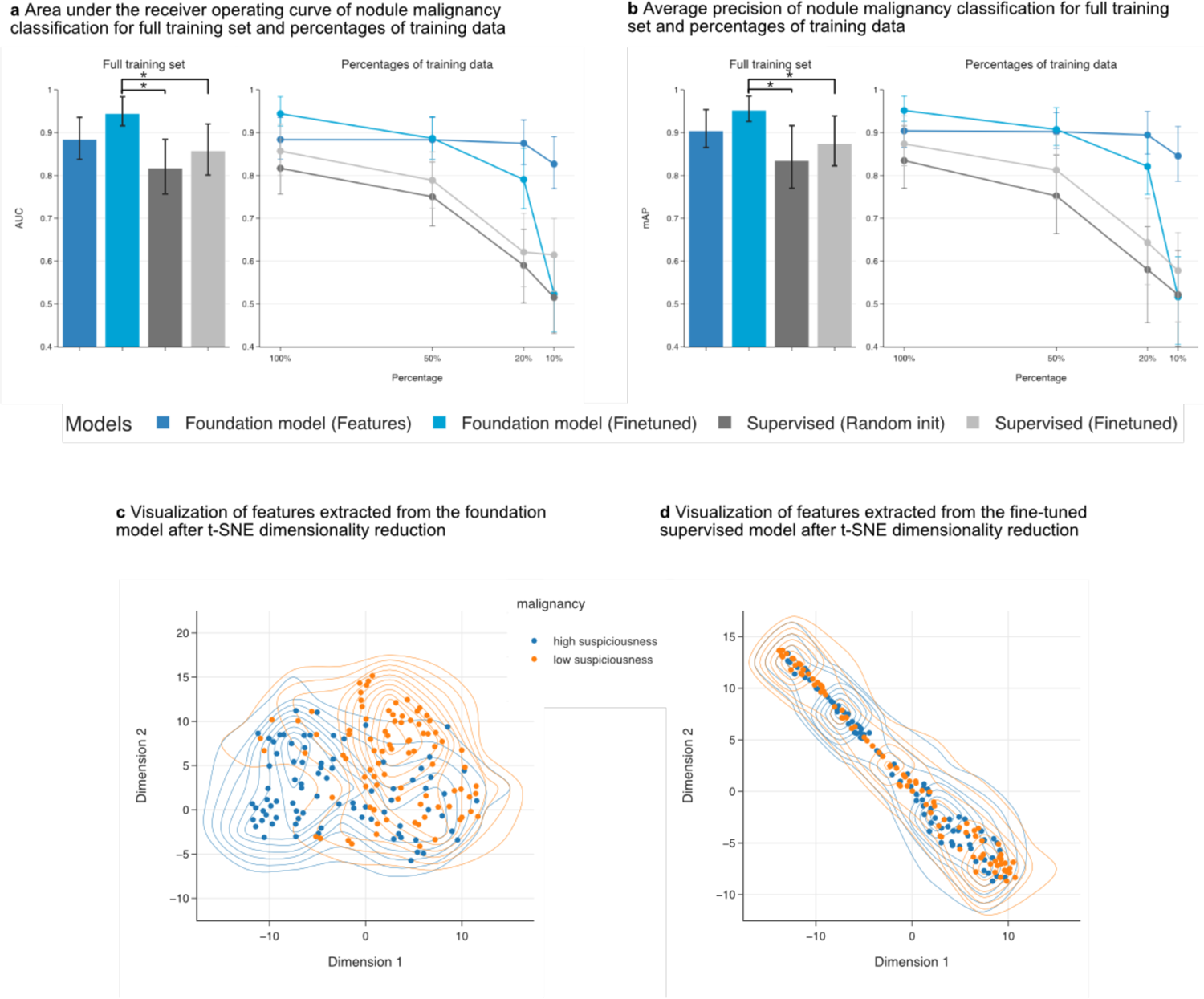
Performance comparison of the foundation model against supervised for nodule malignancy prediction. We compared the foundation model adaptation approaches against baseline supervised models using the full training dataset and on decreasing the training data percentages to 50%, 20% and 10%. **a** Area under receiver operating curves (AUC-ROC) **b** Average precision (AP). Error bars in (**a**) and (**b**) show 95% confidence intervals of the estimates. Visual representation of the features generated from the independent test-set for the task of nodule malignancy prediction using, **c** the fine-tuned supervised model and **d** using the foundation model as a feature extractor. For (**c)** and (**d**), the x-axis corresponds to dimension 1, and the y-axis to dimension 2 of the t-SNE dimensionality reduction. The density contours belonging to each class are underlaid for (**c**) and (**d**) to highlight separability between classes in the feature space.

When analyzing reduced data sizes, the fine-tuned foundation model significantly (p<0.01) outperformed the fine-tuned supervised model when data was reduced to 50% (n=254) and 20% (n=101). However, it did not significantly improve when data was reduced to 10% (n=51). In contrast, the foundation model as a feature-extractor improved significantly (p < 0.005) over all other models at 10%. Moreover, performance from the foundation model as a feature extractor remained relatively stable even when trained on 10% of the data, while all other models showed a significant drop in performance. Across the limited data evaluation, although fine-tuned supervised models showed a trend of improvement over randomly initialized supervised models, they were not found to be significant (p>0.05). Detailed comparisons can be found in the supplementary material (see **Extended Data Table 2**)

We observed that representations from the foundation model demonstrated superior linear discrimination compared to the supervised model, where samples remained interspersed between the classes (see **Fig. 3c, 3d**).

### Prognostication performance for non-small cell lung cancer (NSCLC) tumors (Use case 3)

In the last use case, we evaluated the ability of the foundation model to capture quantitative radiographic phenotypes of NSCLC tumors and consequently determine the prognosis of patients using three independent cohorts of patients treated with surgery or radiation, HarvardRT (n=291), LUNG1 (n=421) and RADIO (n=144) (see use-case 3 in **Fig. 1b**). We aimed to investigate the performance of foundation model implementations when trained and applied to cohorts with strong distribution shifts (cohorts from separate institutions with different standards of care). Therefore, we trained and tuned our prognostication models using data from the HarvardRT cohort to predict 2-year overall survival after treatment and then compared the performance of the foundation model and supervised approaches on the LUNG1 and RADIO cohorts.

In the LUNG1 cohort, foundation models outperformed both supervised methods, with statistical significance (p<0.05). Features extracted from the foundation model obtained an AUC of 0.637 (95% CI 0.583-0.691), and fine-tuning the foundation model resulted in an AUC of 0.619 (95% CI 0.564-0.674), as shown in **Fig. 4a**. In comparison, training supervised models with randomly initialized weights resulted in an AUC of 0.531 (95% CI 0.475-0.587). Fine-tuning a supervised model trained on a different task (use-case 1) showed an AUC of 0.566 (95% CI 0.510-0.622). The best-supervised model (supervised fine-tuned) and the foundation model (features + linear classifier) were evaluated using Kaplan-Meier survival analysis, shown in **Fig. 4c** and **4e**, respectively. The foundation model demonstrated higher prognostic power by better stratifying mortality, as shown by a lower p-value (p<0.0001) when split by the median on the tuning set, compared to the supervised model (p=0.03). Kaplan-Meier curves and univariate Cox regression for all of the models can be found in the supplementary (see **Extended Data** Fig. 1, **Table 3**)

**Figure 4.**
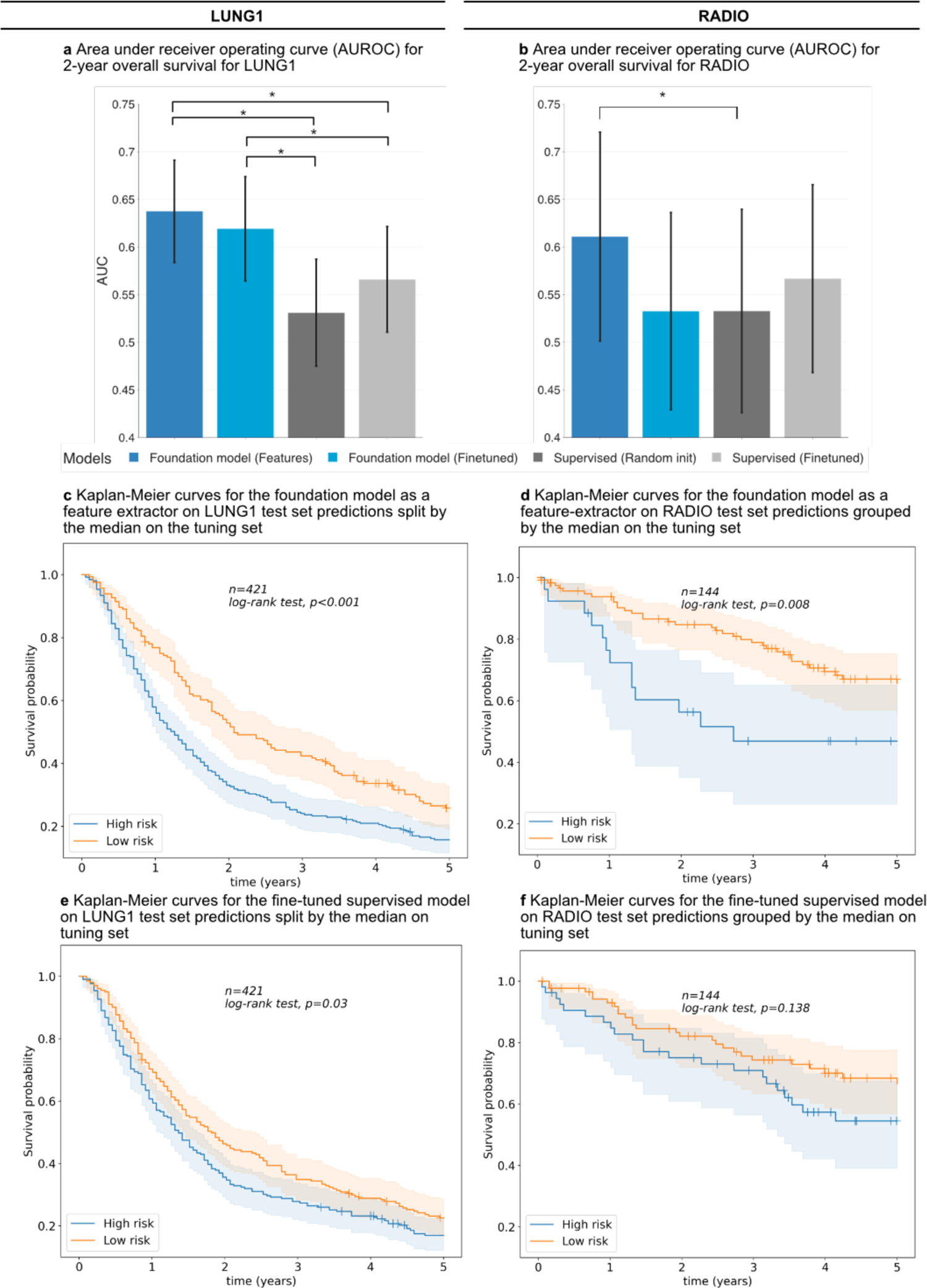
Performance of the foundation model against supervised for prognostication of NSCLC tumors. We compared the foundation model against the baseline supervised models using the area under the receiver operating curve (AUC) for 2-year overall survival for **a** LUNG1 **b** RADIO. Kaplan-Meier (KM) curves for predictions generated from the foundation model as a feature-extractor for LUNG1 (**c)** and RADIO (**d)** as well as the fine-tuned supervised method for LUNG1 (**e**) and RADIO (**f**). To ensure a fair comparison, we calculated the threshold for the split between the KM groups on the tuning set for each network. Kaplan-Meier curves for the other approaches, fine-tuning the foundation model and training a supervised model from random initialization can be found in Fig. S1 in the supplementary.

In the RADIO cohort, the foundation model as a feature extractor performed the best, with an AUC of 0.61 (95% CI 0.501-0.720). Supervised models trained with random initialization had an AUC of 0.532 (95% CI 0.426-0.639) while fine-tuning a supervised model led to an AUC of 0.567 (95% CI 0.468-0.665). Fine-tuning the foundation model did not improve performance, yielding an AUC of 0.532 (95% CI 0.428-0.636), as shown in **Fig. 4b**. Using foundation model features was significantly better than the randomly initialized supervised model (p<0.05), but none of the other networks showed significant differences from the rest (p>0.05). Kaplan-Meier survival analysis demonstrated significant stratification for the feature-extractor foundation model predictions (p=0.008) compared to the fine-tuned supervised model (p=0.138), as shown in **Fig. 4d** and **4f**. Kaplan-Meier curves and univariate Cox regression for all of the models can be found in the supplementary material (see **Extended Data Fig. 1, Table 3**).

### Stability of the foundation model

We evaluated the stability of our foundation model and compared it against supervised approaches in two ways: through a test-retest scenario and an inter-reader variability analysis. To assess test-retest robustness, we used scans from 26 patients from the RIDER dataset^15^ taken within a 15-minute interval using the same imaging protocol. We found that predictions from the best-performing models, feature-extractor foundation, and fine-tuned supervised had high stability with intraclass correlation coefficient (ICC) values of 0.98 and 0.97, respectively. Furthermore, the test-retest features for both networks were strongly correlated (as shown in **Extended Data Fig. 2a and 2b**).

To evaluate stability against inter-reader variability, we used the LUNG1 dataset and perturbed the input seed point to extract the 3D volume, simulating variations among human readers. We found that the feature-extractor foundation models had higher stability against simulated inter-reader variations in prediction performance than the fine-tuned supervised models (see **Extended Data** Fig. 2c and 2d).

### Saliency maps for fine-tuned foundation models

To gain insight into the regions of the input volumes that contribute to a given prediction, we employed gradient-based saliency maps for foundation models fine-tuned on three selected use cases (as depicted in **Fig. 5**). We used smooth guided back-propagation^16,17^ to compute the gradient of the output with respect to the input while keeping the model weights constant. This provided insight into the regions of the input that had the most significant influence on the output prediction.

**Figure 5.**
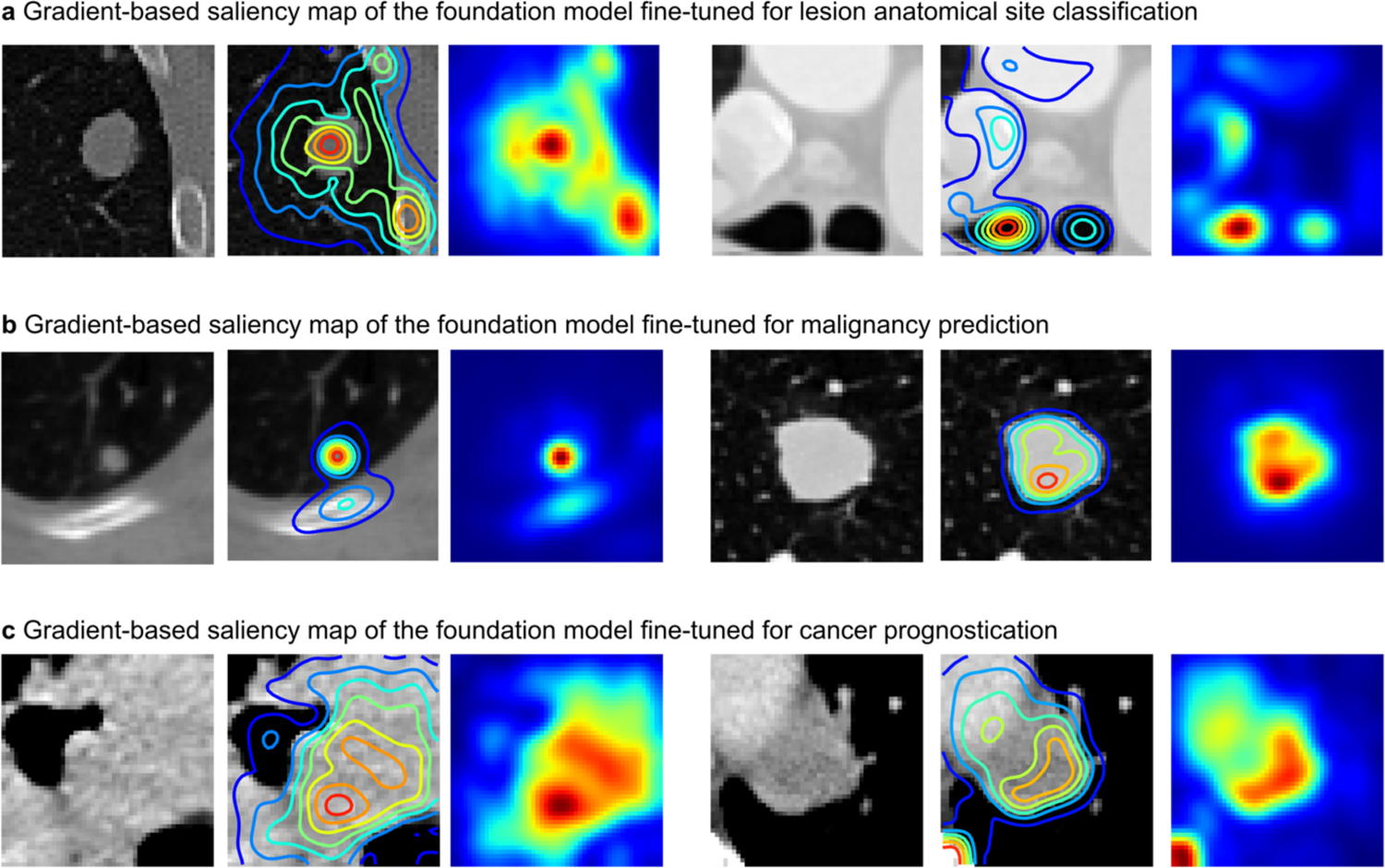
Saliency maps for fine-tuned foundation models. We generated gradient-based saliency maps for each of the fine-tuned foundation models from use cases I (**a**), II (**b**), and III (**c**) using smooth guided backpropagation and visualized salient regions on two samples from corresponding test datasets. The first and fourth columns show the central axial slice (50mm x 50mm) of the volume provided as input to the self-supervised network. The second and fifth columns show isolines for saliency contours. Finally, the third and sixth columns show saliency maps highlighting areas of the input volume that contribute the most to a change in the output prediction.

**Figure 6.**
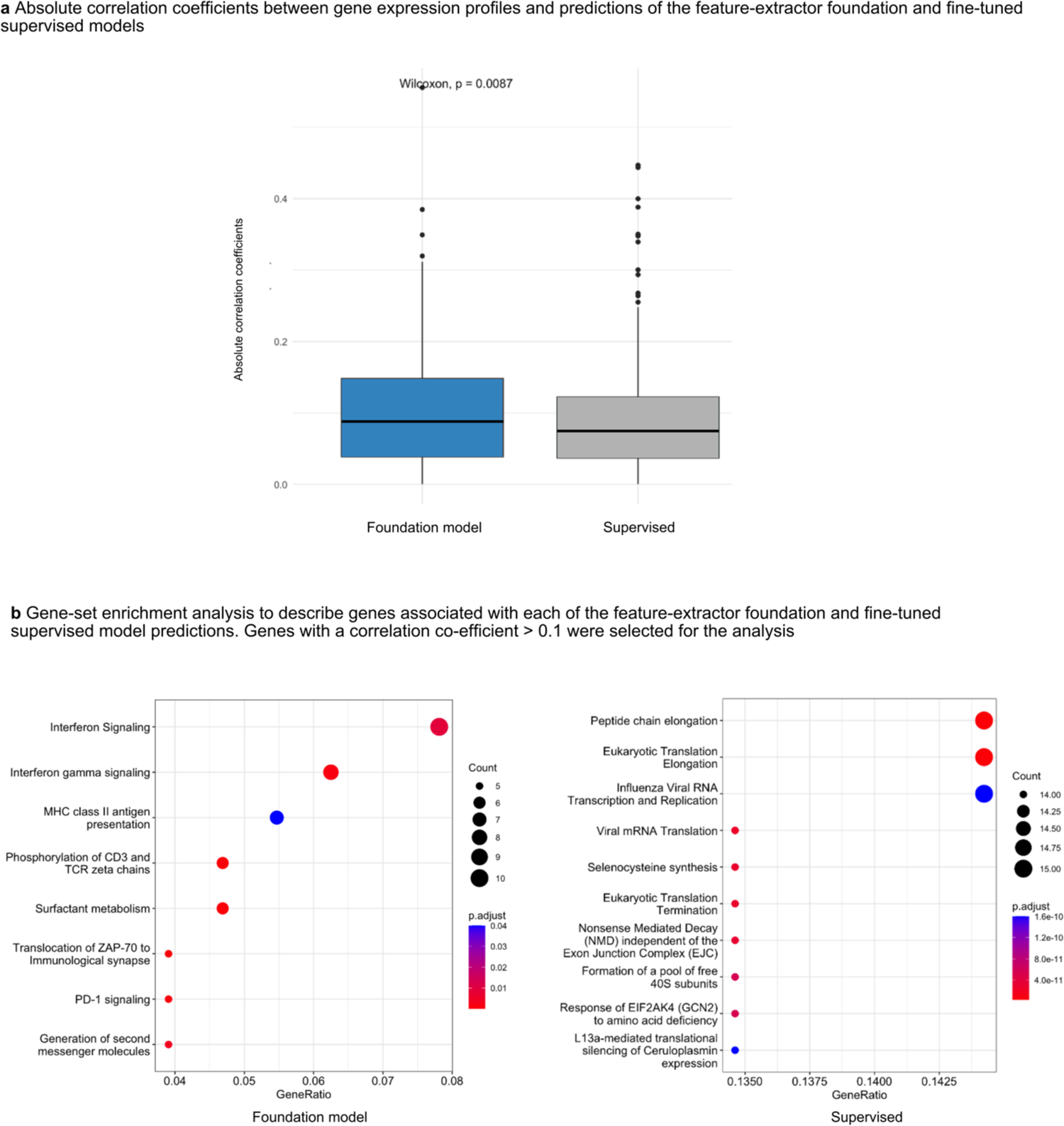
Underlying biological basis of the foundation model. We compared the foundation and supervised model predictions with gene expression profiles. **a** Box plot of absolute correlation coefficients (y-axis) of selected genes against model predictions (x-axis). **b** Gene-set enrichment analysis of genes with correlation coefficient > 0.1 revealed for the foundation (left) and supervised model predictions (right). Genetic pathways are shown on the y-axis, and the gene ratio is shown on the x-axis. Gene count and adjusted p-values are also shown in the legend.

Our analysis revealed that fine-tuned foundation models for each use case focused on different regions but largely converged on tissues within or in proximity to the tumor. This is consistent with research demonstrating the tumor microenvironment’s influence on cancer development^18^ and prognosis. Specifically, lesion anatomical site classification models (as depicted in **Fig. 5a**) focused mainly on areas surrounding the lesions, such as the parenchyma and bone regions in the lung and the trachea in mediastinal lesions. On the other hand, nodule malignancy models (as depicted in **Fig. 5b**) primarily concentrated on the tissues of the nodule while avoiding high-density bone regions. In the case of prognosis networks (as depicted in **Fig. 5c**), the model predictions were primarily attributed to areas surrounding the center of mass of the tumor, with some contribution from high-density bone regions. Overall, these findings indicated that the areas that contribute to the networks’ predictions varied in accordance with the specific use case, with the tumor and surrounding tissues playing a pivotal role.

### Underlying biological basis of the foundation model

Finally, we investigated the biological basis of our foundation model by analyzing gene expression data associated with model predictions for 130 subjects from the RADIO dataset. To identify relevant genes, we selected the top 500 genes and performed a correlation analysis, comparing the feature-extractor foundation and fine-tuned supervised model predictions with gene expression profiles. We found that absolute correlation coefficients between gene expression profiles and model predictions were significantly higher (p=0.008) for the foundation model, indicating a stronger association with underlying tumor biology (see **Fig. 6a**).

Additionally, we examined the genes associated with these models through a gene set enrichment analysis (genes with a correlation coefficient> 0.1). Our analysis revealed that foundation models showed a pattern of enrichment of immune-associated pathways, including interferon signaling, interferon gamma signaling, MHC class II antigen presentation, and PD-1 signaling. Conversely, while the supervised model did show enrichment of individual pathways, no identifiable pattern was observed (see **Fig. 6b**).

## DISCUSSION

In this study, we demonstrated that our foundation model trained using self-supervised learning, provided robust quantitative biomarkers for predicting anatomical site, malignancy, and prognosis across three different use cases in four cohorts. Several studies^19–21^ have demonstrated the efficacy of self-supervised learning in medicine where only limited data might be available for training deep learning networks. Our findings complement and extend this for identifying reliable imaging biomarkers for cancer-associated use cases. We showed that our foundation model provided superior performance for anatomical lesion site and malignancy prediction. Modeling using features extracted from the foundation model was the most robust across tasks offering stable performance even when data sizes were considerably reduced to 51 samples (10% of use-case 2). These features could also categorize data from these tasks into semantically separable clusters corresponding strongly with target classes, although these features were learned independent of class information. Using these features provided the best performance on small cohorts in predicting prognosis and also demonstrated significant stratification of patients by their associated risk for each of the LUNG1 and RADIO cohorts (p<0.01). Additionally, predictions using the foundation model features were found to be highly stable against inter-reader (standard deviation=0.004) and test-retest variations (ICC=0.98). Regarding the interpretability of features, we observed that models focused on varying regions of the tumor and surrounding tissue relevant to the associated use case. To gain insight into the underlying biological associations of these features, RNA sequencing analysis combined with imaging data showed that these features correlated with immune-associated pathways.

Studies for predicting endpoints, such as overall survival on small cohorts largely rely on statistical feature extraction (engineered radiomics) and classical machine learning-based modeling. Precise three-dimensional segmentations are required for extracting these statistical features from tumor volumes increasing the annotation burden associated with these studies. Moreover, these statistical features are affected by several confounders, such as inter-reader variability in segmentations ^22^ and acquisition settings of the scanners ^23^. Deep learning methods, in comparison, are robust to differences in acquisition and segmentation variability and provide improved performance over statistical features ^10^. However, they remain restricted in their applicability in such low-data scenarios due to their dependency on large amounts of data to provide robust performance. Training deep-learning models on small cohorts often lead to overfitting, which diminishes performance when external data is introduced^11^. Our foundation model approach has several innovations: first, we developed a deep-learning system on a large corpus of 3D lesion images with considerable diversity in their presentation. To our knowledge, our study is the first to pre-train a deep-learning model using 11,467 3-dimensional lesion volumes. Second, we demonstrated that our pre-trained model learned generalizable features and improved performance across three tasks and associated endpoints. Our model also provided prognostic value when trained on small cohorts and applied to external validation cohorts. Third, our models showed high robustness to test-retest and inter-reader variations. Finally, we share our validated foundation model with the public, allowing external testing and future studies to facilitate their adoption into external workflows.

Several studies have investigated deep learning algorithms for identifying cancer imaging biomarkers in both small and large cohorts. Hosny et al.^10^ trained a deep learning model for lung cancer prognostication using several multi-institutional cohorts and demonstrated strong performance using deep learning methods over traditional radiomics features. Kumar et al.^24^ identified radiomic sequences using deep convolutional encoders for determining the malignancy of lung nodules from the LIDC-IDRI dataset considering 4306 lesions. Lao et al.^25^ proposed a deep-learning model-based radiomics signature for predicting survival in glioblastoma multiforme, trained and validated on relatively small cohorts. Haarburger et al.^26^ present a deep convolutional network-based approach to predict survival endpoints on the LUNG1 dataset. Cho et al.^27^ developed a radiomics-guided deep-learning model for stratifying the prognosis of lung adenocarcinoma and validated it in a local cohort and an external validation cohort. A general trend observed across these studies is that the performance of deep learning models is more robust when larger and multi-institutional cohorts are available for training. Validation is subsequently performed on cohorts smaller than the training cohort. A demonstrated strength of our approach is that training on smaller cohorts performs well in larger validation cohorts. For the prognostication use case, we performed well on two external validation cohorts with a combined size considerably larger than the training cohort. Our pre-trained foundation model shows strong generalization ability across our diverse use cases and may apply to several other cancer imaging use cases out of the box. Furthermore, extracting features from our model (inference only) followed by simple modeling methods is resource-efficient, alleviating the need for expensive hardware for training standard deep-learning models while providing on-par performance.

In recent years, self-supervised pre-training has been applied to medical imaging with promising results^19,21,28,29^. Zhou et al.^30^ present an approach that constructs several pre-text tasks to train SSL networks and show that they outperform solely supervised networks trained across five clinically relevant tasks. A novel contrastive SSL strategy incorporating both global and local information captured within medical images and reporting their superior performance, especially in low-data settings, is proposed by Chaitanya et al.^31^. Azizi et al.^19^ demonstrate that grouping multiple images attributed to the same medical condition along with combining natural and medical images for contrastive SSL training improves performance. Specifically for deep radiomics applications, Li et al.^32^ propose targeting data imbalance in existing data and present a combined approach of traditional radiomic features and self-supervised learning representations, improving performance for discriminating tumor grade and tumor staging tasks. Li et al.^33^ proposed a novel self-supervised collaborative approach for creating latent representations from radiomic features. Zhao and Yang^34^ used self-supervised learning to pre-train models via a radiomic-deep feature correspondence task. Although these studies have investigated self-supervised learning for radiomics tasks, they lacked external validation or proposed limited evaluation of the generalizability of their approaches. Our study presents a foundation model for radiomic discovery by pre-training on a large cohort of lesions. The examined tasks are independent of the pre-training cohort and demonstrate the increased generalizability of our proposed approach.

Despite the strengths outlined in our study, we recognize several limitations that need to be addressed prior to the clinical applicability of our foundation model. Features from the foundation model followed by linear classifiers provided the most robust performance across all investigated tasks. However, linear classifiers might be sub-optimal in identifying complex relationships between feature representations to predict challenging endpoints. As we aimed to demonstrate the benefits of our foundation model compared to existing approaches, we have limited our exploration with fine-grained feature and model selection strategies. Comprehensive selection approaches similar to Parmar et al.^35^ might improve performance even further, strengthening our hypothesis for foundation models.

Similarly, deep learning-based finetuning approaches employed in this study are representative of baseline performance. We observed that finetuning approaches for the foundation model in low data settings (especially 10%) and smaller cohorts (HarvardRT) resulted in suboptimal performance compared to using extracted features. We hypothesize that in lower data settings, models overfit the training data and demonstrate worse generalization as the number of parameters to tune increases. However, with the steady emergence of deep learning literature proposing improvements to handle aspects such as data imbalance, hyperparameter selection, and optimization objectives, the performance of these models can be pushed far above the current baseline. Our prognostication model is also limited in its performance due to our focus on solely imaging data; incorporating clinical features has a large potential to improve its effectiveness.

Our foundation model’s clinical applicability encounters challenges typically associated with deep learning, including generalizability, interpretability, and explainability. Given the retrospective nature of this study, our capacity to evaluate the real-world practicality of foundation model-based biomarkers is constrained. Deep learning models are notorious for being black boxes that offer little clarity on interpretable and explainable reasoning behind their predictions. Although we used well-established saliency attribution methods to interpret our foundation model’s predictions, the broader applicability of these insights is hindered by the technical limitations of such methods ^36,37^. In addition to the limitations of deep learning methodology, the biological association analysis conducted to explain our model’s predictions is preliminary and requires further investigation to generate a concrete understanding. We anticipate that future external validation of our open-access model will help confront these prevalent challenges.

In conclusion, our foundation model offers a powerful and reliable framework for discovering cancer imaging biomarkers, even in small datasets. Furthermore, it surpasses current deep learning techniques in various tasks while fitting conveniently into existing radiomic research methods. This approach can potentially uncover new biomarkers that significantly contribute to research and medical practice. We share our foundation model and reproducible workflows so that more studies can investigate our methods, determine their generalizability, and incorporate them into their research studies.

## METHODS

### Study Population

We utilize a total of five distinct datasets, four of which are publicly accessible, and one is an internal dataset. These were acquired from various institutions as components of separate investigations (see **Extended Data Table 4**).

DeepLesion^14^ is a dataset comprising 32,735 lesions from 10,594 studies of 4,427 unique patients collected over two decades from the National Institute of Health Clinical Center PACS server. Various lesions, including kidney, bone, and liver lesions - as well as enlarged lymph nodes and lung nodules, are annotated. The lesions are identified through radiologist-bookmarked RECIST diameters across 32,120 CT slices. In our study, we excluded CT scans with a slice thickness exceeding 3mm, resulting in 16,518 remaining lesions. Subsequently, we divided this into 11,467 unlabelled lesions for contrastive training and 5,051 labeled lesions for anatomical site classification. The labeled lesion data were further separated randomly into training, tuning, and testing sets, containing 2,610, 1,220, and 1,221 lesions, respectively.

LUNA16^38^ is a curated version of the LIDC-IDRI dataset of 888 diagnostic and lung cancer screening thoracic CT scans obtained from seven academic centers and eight medical imaging companies comprising 1,186 nodules. The nodules are accompanied by annotations agreed upon by at least 3 out of 4 radiologists. Alongside nodule location annotations, radiologists also noted various observed attributes like internal composition, calcification, malignancy, suspiciousness, and more. For our evaluation, we chose nodules with at least one indication of malignancy suspicion, totaling 677. We randomly picked 338 nodules for training and 169 for tuning the malignancy prediction networks. The final 170 nodules were utilized to assess the networks’ performance.

HarvardRT^10^ is a cohort of 317 patients with stage I-IIIB NSCLC treated with radiation therapy at the Dana-Farber Cancer Institute and Brigham and Women’s Hospital, Boston, MA, US, between 2001 and 2015. All CT scans for this cohort were acquired with and without intravenous contrast on the GE Lightspeed CT scanner. The primary tumor site was contoured by radiation oncologists using soft tissue and lung windows. A subset of 291 patients with a follow-up of 2 years was selected for this study. We used 203 tumor volumes for training the prognostication networks and the remaining 88 tumor volumes for tuning.

LUNG1^39^ is a cohort of 422 patients with stage I-IIIB NSCLC treated with radiation therapy at MAASTRO Clinic, Maastricht, The Netherlands. FDG PET-CT scans were acquired with or without contrast on the Siemens Biograph Scanner. Radiation oncologists used PET and CT images to delineate the gross tumor volume. For our study, we selected CT scans of 421 patients with annotated primary gross tumor volumes and used these as an independent test set for prognostication networks.

RADIO (NSCLC-Radiogenomics)^40^ dataset is a collection of 211 NSCLC stage I-IV patients recruited between 2008 and 2012 who were referred for surgical treatment and underwent preoperative CT and PET/CT scans. These patients were recruited from the Stanford University School of Medicine and the Palo Alto Veterans Affairs Healthcare System. Scan scans were obtained using various scanners and protocols depending on the institution and physician. A subset of 144 patients in the cohort has available tumor segmentations independently reviewed by two thoracic radiologists. In addition to imaging data, the dataset includes molecular data from EGFR, KRAS, ALK mutational testing, gene expression microarrays, and RNA sequencing. For the current study, we utilized the subset of 144 patients with annotated gross tumor volumes as an independent test set for prognostication and also investigated the biological basis of our networks using this dataset.

### Data Preprocessing

CT scans were resampled using linear interpolation to achieve isotropic voxels with a 1mm³ resolution to address variations in slice-thickness and in-plane resolutions across study populations. We extracted patches of 50 x 50 x 50 voxels from the scans centered around a seed point (refer to **Extended Data** Fig. 3). For the DeepLesion dataset, which provided annotations in the form of RECIST diameters, the seed point was determined by calculating the midpoint of the RECIST diameter. For the other datasets (i.e., LUNA16, HarvardRT, LUNG1, and RADIO), which supplied annotations as 3D contours, the seed point was obtained by computing the center of mass (CoM). This approach allows for significantly higher throughput than manual segmentation, which can be more tedious. We then normalized the voxel values in the patches by subtracting −1024 (lower-bound Hounsfield unit) and dividing by 3072 (upper-bound Hounsfield unit), ensuring the intensity values in the input data ranged between 0 and 1.

### Task-agnostic contrastive pre-training of the foundation model

We implemented contrastive pre-training using a modified version of the SimCLR framework^5^. The SimCLR framework’s general principle involves transforming a single data piece (e.g., a patch taken from a CT scan) into two correlated and augmented samples (e.g., the same patch rotated 15 degrees clockwise and flipped horizontally). A convolutional encoder is then used to extract latent representations from these samples. Through a contrastive loss function^41^, the model learns to identify similar representations from the same data sample and dissimilar representations from different data samples. The framework emphasizes effective transformation choices, convolutional encoder architectures, and contrastive loss functions for optimal self-supervised learning performance. To effectively represent the nature of medical images, we made modifications to each of these components.

Transformations proposed in the original SimCLR framework for natural world images, such as cutout augmentation, Sobel filtering, and color distortion, are unsuited for 3D medical images due to dynamic range and color depth differences. Therefore, our study applies different augmentations to replace these transformations. For instance, we substituted the random color jitter transform with a random histogram intensity shift transform, as they both induce variation in intensity distribution.

To extract representations from the transformed 3D volumes, we selected the 3D ResNet50 architecture as our deep convolutional encoder. While the SimCLR authors employed a 2D ResNet50 architecture, we opted for its 3D counterpart, which has proven effective in handling 3D medical imaging data^42^.

Regarding loss functions, we extended normalized temperature-scaled cross-entropy loss (NT-Xent)^43^ to support contrastive training for lesion volumes. The modifications include: 1) selecting positive pairs as 3D patches surrounding the lesion’s seed point, 2) choosing negative pairs by randomly sampling 3D patches from the rest of the scan, and 3) computing the contrastive loss on these positive and negative pairs, with each iteration comprising N positive pairs and N*2(N-1) negative pairs. We also explored different temperature parameters for the NT-Xent loss. However, the original value of 0.1 proposed by the original paper was the most effective.

Our model was pre-trained for 100 epochs using an effective batch size of 64 (32 x 2 training nodes) on two NVIDIA Quadro RTX 8000 GPUs taking approximately five days. We used Stochastic Gradient Descent (SGD) as the optimizer, with layer-wise adaptive rate control (LARC), momentum, and weight-decay enabled. To improve the optimization process, we employed learning rate schedulers that combined linear and cosine decay strategies and a warmup phase to modify the learning rate at the beginning of training gradually. While most specifications were consistent with the original SimCLR experiments, we experimented with different batch sizes, patch sizes (50mm³ and 64mm³), learning rates, transforms, and model architectures.

### Task-specific training of the foundation model

Our foundation model was adapted for a specific task through two approaches: 1) extracting features and fitting a linear classifier on top of them or 2) fine-tuning the pre-trained ResNet50 for the given classification task.

We extracted 4096 features from the foundation model for each data point and used them to train a logistic regression model using the scikit-learn framework^44^. A comprehensive parameter search for the logistic regression model was performed using the optuna hyper-parameter optimization framework^45^. No performance improvements were observed through feature selection strategies; therefore, all 4096 features were used in accordance with linear evaluation strategies prevalent in self-supervised learning (SSL) literature.

Fine-tuning was carried out with all layers updated during training, utilizing cross-entropy loss. A series of randomly chosen augmentations—random flips, random 90-degree rotations, and random translations of ±10 voxels across all axes—were applied throughout the training. SGD was employed for network training, with momentum enabled and step-wise learning rate decay. Following the original SimCLR experiments, configurations and similar parameters (including learning rate, transforms, and model architectures) were explored during hyperparameter tuning. Each network was trained for 100 epochs using a single NVIDIA Quadro RTX 8000 GPU, and the best-performing model checkpoints was chosen based on the tuning set.

For supervised baseline models, their weights were initialized randomly, and they were trained using the same configuration that was adopted for fine-tuning the foundation model. The supervised models for use cases 2 and 3 were also fine-tuned, utilizing the same configuration as in the pre-trained fine-tuning process but by initializing them with the weights of the trained supervised baseline from use case 1.

Task-specific training was conducted on reduced dataset sizes in addition to utilizing the entire dataset. We randomly sampled 50%, 20%, and 10% of the training and tuning datasets and constructed task-specific models using these samples with the same configuration as the entire dataset. As the training dataset sizes decreased, we considered training the models for a higher number of epochs; however, models frequently overfitted during extended training. The entire test dataset was employed to allow benchmarking across these splits.

### Performance Analysis

Validation of the foundation model was performed using several use-case-relevant metrics. Lesion anatomical site classification performance was assessed using balanced accuracy (BA) as a multi-label counting metric and mean average precision (mAP) as a multi-threshold metric. The multi-label metric, BA, adjusts class-wise accuracy based on the class distribution at a chosen threshold (0.5). The multi-threshold metric, mAP, enables the examination of a given class’s performance across a range of prediction thresholds. All classes other than the class of interest are considered negatives, and performance is averaged across all possible classes. We avoided using the area under the receiver operating curve (AUC-ROC) for this use case due to the high proportion of negatives relative to positives, which results in consistently low false-positive rates and might overestimate the AUC. However, due to a more balanced class distribution, nodule malignancy prediction was evaluated using AUC-ROC. NSCLC prognostication networks also employed AUC-ROC for evaluation, as it estimates the ranking of subjects based on their survival times.

Models underwent pairwise comparison using permutation tests. N permutations (N=1000) were conducted for each pair, and new models were computed after permuting class labels. Metrics were recalculated after resampling, and a two-sided p-value was calculated to test the null hypothesis of observations from each pair originating from the same underlying distribution. Additionally, 95% confidence intervals were established for each model using a bootstrap test with N=9999 resamples.

Kaplan-Meier (KM) curves were also used to determine the stratification of subjects based on their prediction scores for the prognostication models. Groups were selected based on prediction scores on the tuning set, and curves were plotted on the test set for these groups. Multivariate log-rank tests were used to examine the significance of the stratification. Univariate Cox regression models were built using the model predictions as the categorical variables of interest, grouped similarly to the KM curve.

### Feature visualization and saliency maps

We used the foundation and top-performing supervised models as feature extractors to obtain 4096 distinct features per data point. To enable visual interpretation of these high-dimensional features, we utilized t-SNE^46^ (t-Stochastic Neighbourhood Embeddings) to reduce their dimensionality to 2D. To arrive at the most interpretable visualization, we explored various parameter configurations, including perplexity, initialization, and learning rates. Points in the 2D visualization were color-coded according to their respective target classes, despite dimensionality reduction being agnostic to these distinctions. Density contours were superimposed over the visualizations to enhance the understanding of group patterns, offering a more comprehensive representation of trends across data points.

In order to generate saliency maps for each task, the fine-tuned foundation model was used to generate predictions on randomly selected volumes from respective datasets. The fine-tuned foundation model with a single output prediction (corresponding to the predicted target class) was chosen in contrast to the feature extractor as computing saliency maps over 4096-dimensional outputs remains challenging in practice. We used a combination of 1) smooth gradient backpropagation, which averages gradients of the output with respect to several noisy inputs, and 2) guided back-propagation which combines deconvolution with backpropagation, mainly stopping the flow of negative gradients or neurons that decrease the activation signal. The method is termed smooth guided-backpropagation and is implemented in the MONAI framework^47^.

### Stability Testing

To test the stability of our models, we performed a test-retest stability and inter-reader variation evaluation. For the test-retest evaluation, we compared model predictions (of outcome) from the best foundation and supervised models generated on chest CT scans taken in a 15-minute interval for 32 patients. Intraclass correlation coefficient (ICC) was computed using the interrater reliability and agreement package (*irr*) in R^48^. We also tested the stability of the flattened features computed by the models by calculating Spearman correlation and R^2^.

For the inter-reader variation evaluation, we used the LUNG1 dataset and generated 50 random perturbations sampled from a three-dimensional multivariate normal distribution with zero mean and diagonal covariance matrix for each seed point. Across each dimension, a variance of 16 voxels was used for generating samples. We generated predictions on perturbed seed points using the best foundation and supervised model, resulting in 50 different prediction models for each. The mean and variance of the 50 models were computed for each and compared.

### Biological Associations

The GSE103584 dataset contains 130 NSCLC (Non-Small Cell Lung Cancer) samples that consist of paired CT scans and gene expression profiles generated by RNA sequencing. To analyze gene expression profiles, we filtered them based on cohort mean expression and standard deviation. First, we took only the genes with a higher expression than the overall dataset mean and then picked the top 500 genes based on standard deviation. Next, we performed a correlation analysis comparing the best-supervised and foundation models. To further evaluate foundation model features’ association with tumor biology, we computed the absolute value of the correlation coefficients and performed a gene set enrichment analysis with all genes with a correlation coefficient above 0.1.

## Data Availability

The majority of the datasets utilized in this study are openly accessible for both training and validation purposes and can be obtained from the following sources: i) DeepLesion [https://nihcc.app.box.com/v/DeepLesion], used both for our pre-training and use-case 1 ii) LUNA16 [https://luna16.grand-challenge.org] used for developing our diagnostic image biomarker iii) LUNG1 [wiki.cancerimagingarchive.net/display/Public/NSCLC-Radiomics] and iv) RADIO [https://wiki.cancerimagingarchive.net/display/Public/NSCLC-Radiomics] used for the validation of our prognostic image biomarker model. The training dataset for our prognostic biomarker model, HarvardRT, is internal and unavailable to the public. HarvardRT was collected under an IRB-approved retrospective protocol with a waiver of consent (Dana-Farber/Harvard Cancer Center protocol 11-286). As the trained foundational model is public, all the results can be reproduced using the accessible test datasets.

https://wiki.cancerimagingarchive.net/display/Public/NSCLC+Radiogenomics

## ACKNOWLEDGEMENTS

The authors acknowledge financial support from NIH (H.J.W.L.A: NIH-USA U24CA194354, NIH-USA U01CA190234, NIH-USA U01CA209414, and NIH-USA R35CA22052), and the European Union - European Research Council (H.J.W.L.A: 866504).

## AUTHOR CONTRIBUTIONS

Study conceptualization: S.P, H.J.W.L.A.; Data acquisition, analysis, and interpretation: S.P, D.B, A.H, T.L.C, H.J.W.L.A.; Methodological design and implementation: S.P, D.B.; Conceptualization of assessment strategies: S.P, D.B, N.J.B, H.J.W.L.A; Statistical Analyses: S.P, M.S, N.J.B, H.J.W.L.A; Code and reproducibility: S.P, I.H, V.P; Writing of the manuscript: S.P, D.B, M.S, S.B, H.J.W.L.A; Critical revision of the manuscript: All authors; Study supervision: H.J.W.L.A

## CODE AVAILABILITY STATEMENT

The complete pipeline used in this study can be accessed either from the <u>AIM webpage</u> or directly on <u>GitHub</u>. This includes the code for 1) Data download and pre-processing: Starting from downloading the data to generating splits used in our study; 2) Replicating the training and inference of foundation and supervised models across all tasks; and 3) Code for reproducing our comprehensive performance validation. In addition to the code, we also provide trained model weights, extracted features, and outcome predictions for all the models used in our study. Most importantly, we provide our foundation model accessible through a simple pip package install and 2 lines of code to extract features for your data. We also provide a detailed documentation website that can be accessed <u>here</u>. The final model weights will also be made available through the Zenodo.org platform as well as through <u>Mhub.ai</u> in a reproducible, containerized, off-the-shelf executable format.

## EXTENDED DATA

**Extended Data Figure 1.**
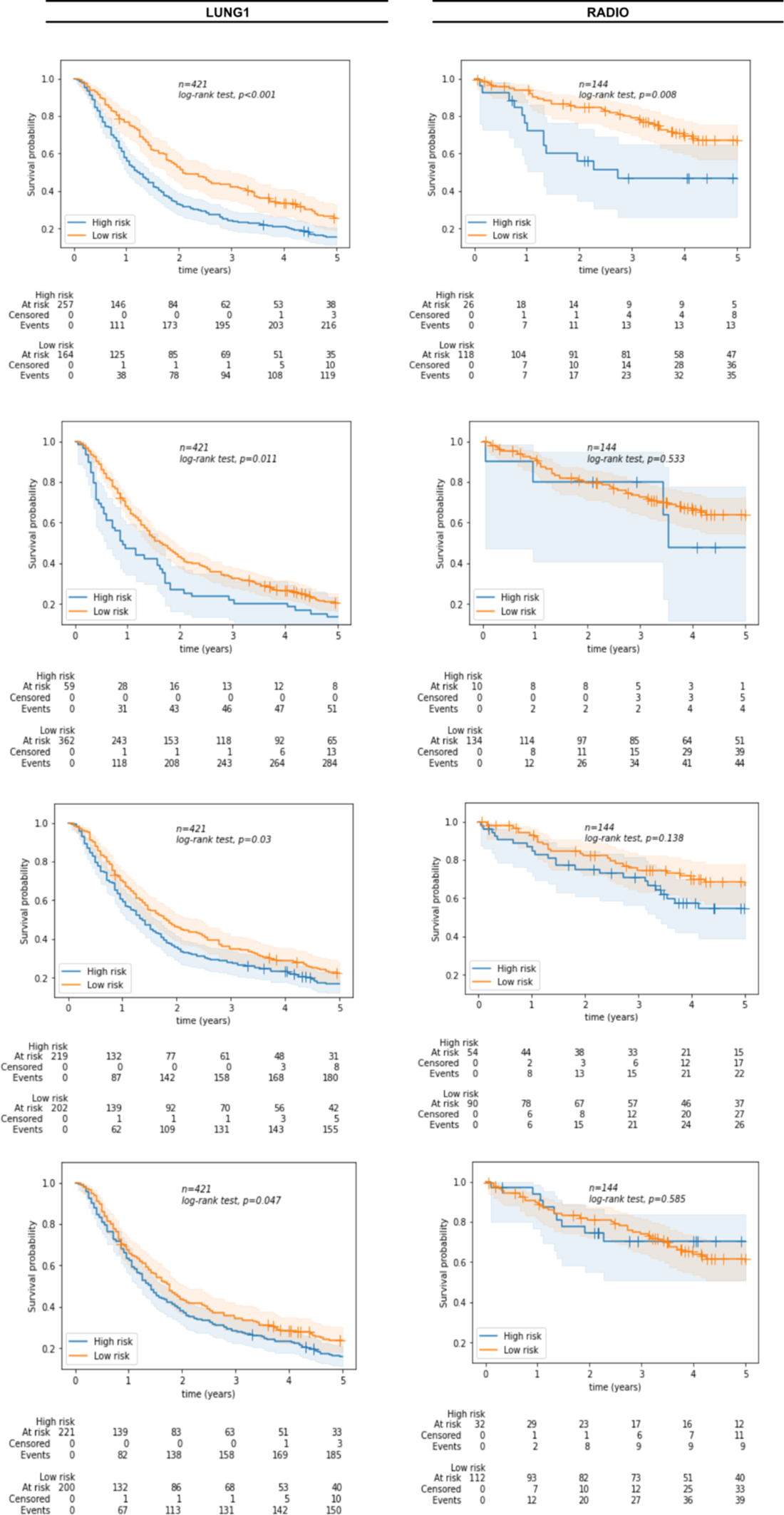
Kaplan Meier curves for all models investigated. Kaplan Meier curves for the LUNG1 and RADIO datasets for the foundation model as a feature-extractor (first row), fine-tuned foundation model (second row), fine-tuned supervised model (third row) and randomly initialised supervised model (last row)

**Extended Data Figure 2.**
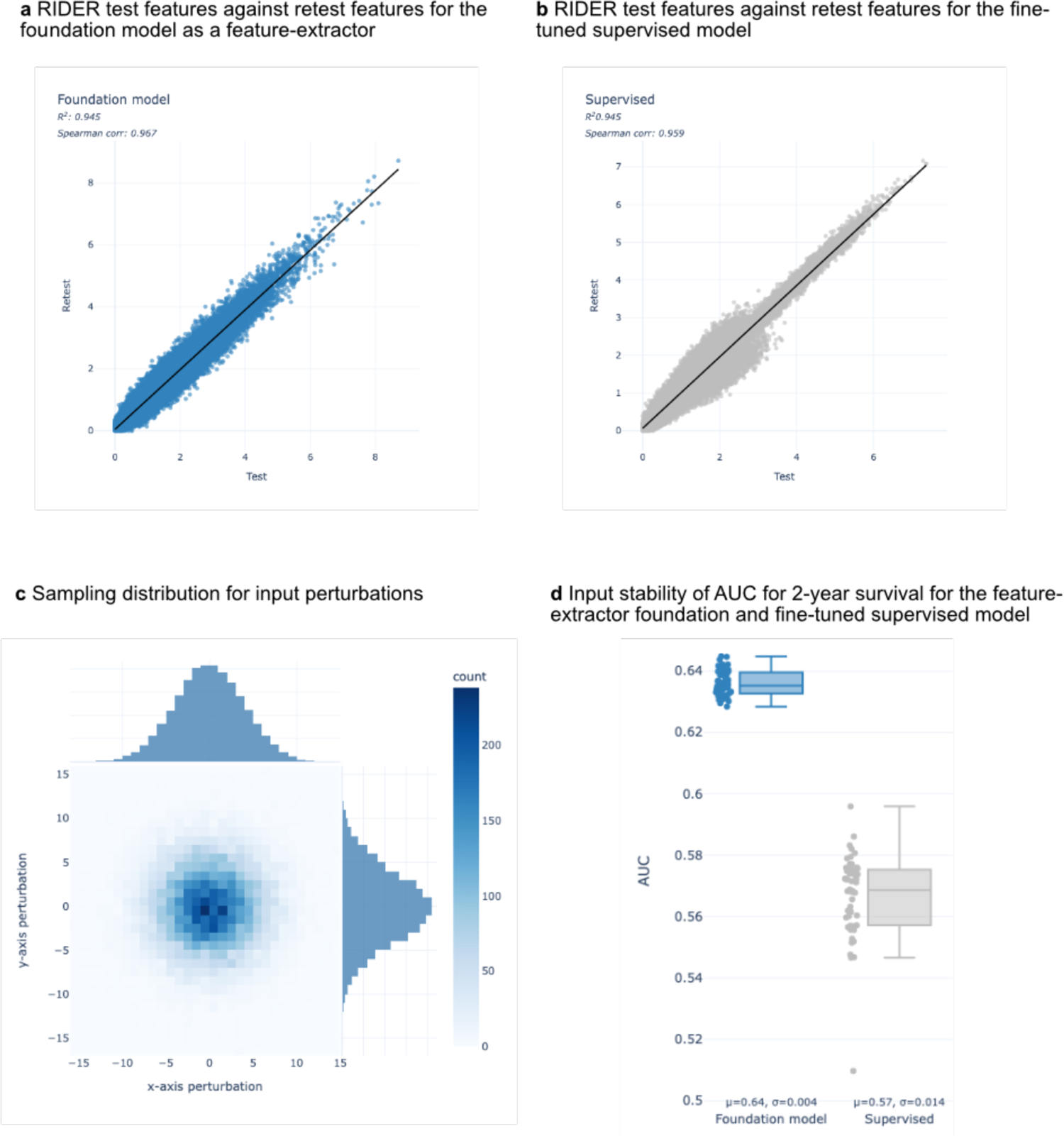
Stability of self-supervised learning networks. We analyzed the test-retest robustness on the RIDER dataset by comparing the correlation between features generated by **a**. the foundation model as a feature extractor and **b.** the fine-tuned supervised model. In **c**., the inter-reader variability is simulated by adding perturbations from a sampling distribution. We perturb across x, y and z-axes although the distribution is shown only for x and y perturbations for simplicity. **d** Prognostic stability of the feature extractor foundation model against the fine-tuned supervised model when the input seed point is perturbed, estimated through AUC for 2-year survival.

**Extended Data Figure 3.**
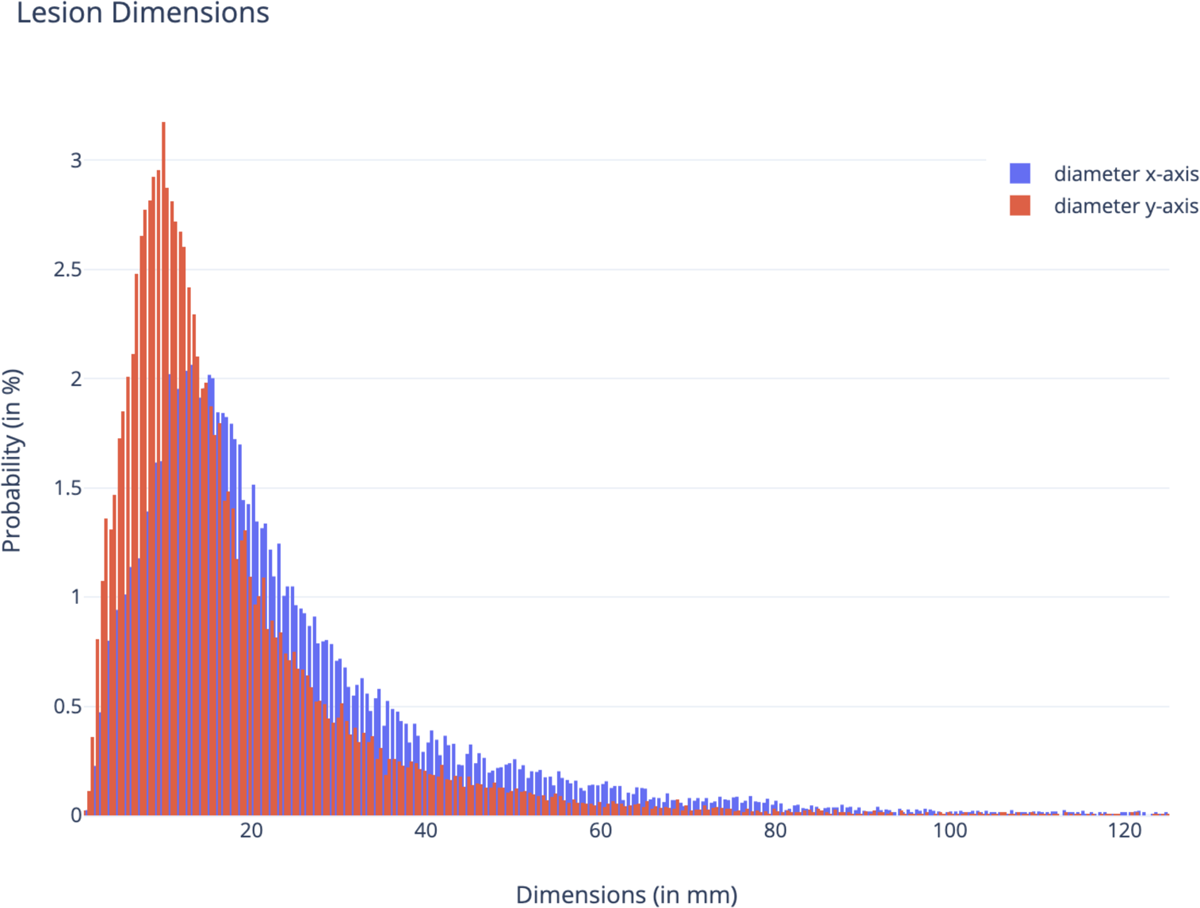
Diameter distribution of DeepLesion. Distribution of diameters in the x and y axes for the DeepLesion training dataset based on RECIST bookmarks identified on key slices. Input dimensions of 50×50×50 mm^3^ were chosen as they covered 93% and 97% of the distribution in the x and y axes respectively.

---

**Extended Data Table 1.** Detailed comparison of the foundation model implementations against supervised methods in limited data settings for lesion anatomical site classification Comparison of the foundation model as a feature-extractor and fine-tuned against the randomly initialised supervised model at 50%, 20% and 10% training data. For each data percentage, the largest increase in performance between the two is shown italicised. Not significant results are shown in red.

| Foundation Model Implementation | Data percentage | Increase in BA over supervised (95% CI, p-value) | Increase in mAP over supervised (95% CI, p-value) |
| --- | --- | --- | --- |
| Feature-extractor | 50%<br>(n=2526) | 0.153 (0.123-0.186,<br>p<0.005) | 0.135<br>(0.104-0.168,<br>p<0.005) |
| Fine-tuned |  | 0.181 (0.147-0.214,<br>p<0.005) | 0.127<br>(0.097-0.162,<br>p<0.005) |
| Feature-extractor | 20%<br>(n=1010) | 0.194 (0.159-0.228,<br>p<0.005) | 0.177<br>(0.142-0.216,<br>p<0.005) |
| Fine-tuned |  | 0.130 (0.102-0.159,<br>p<0.005) | 0.121<br>(0.089-0.159,<br>p<0.005) |
| Feature-extractor | 10%<br>(n=505) | 0.189 (0.148-0.228,<br>p<0.005) | 0.149<br>(0.112-0.189,<br>p<0.005) |
| Fine-tuned |  | 0.063<br>(0.028-0.098, p<0.005) | 0.02<br>(-0.011- 0.061,<br>p=0.28) |

**Extended Data Table 2.**
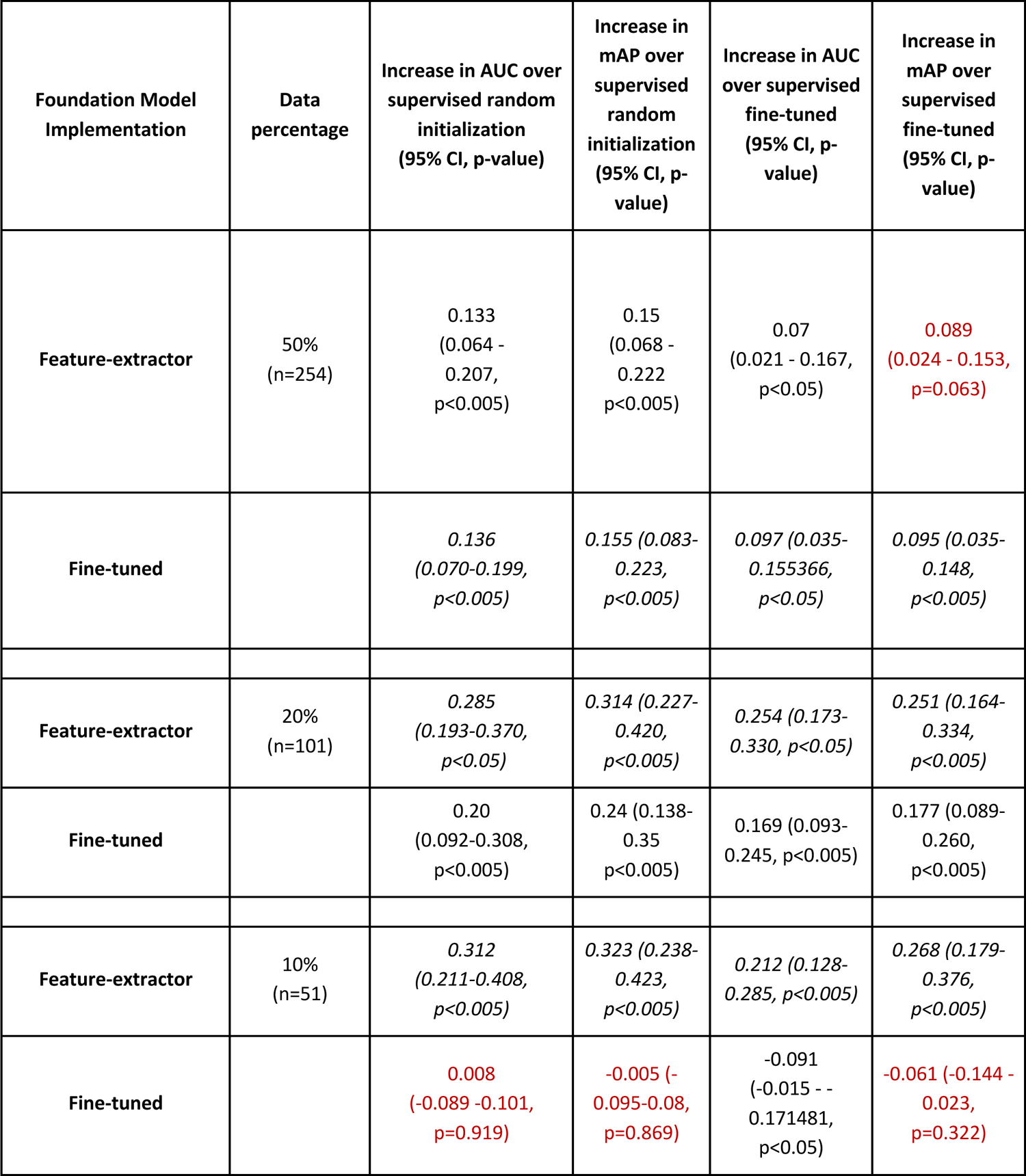
Detailed comparison of the foundation model implementations against supervised methods in limited data settings for nodule malignancy classification Comparison of the foundation model as a feature-extractor and fine-tuned against randomly initialised and fine-tuned supervised models at 50%, 20% and 10% of the training data. For each data percentage, the largest increase in performance between the two is shown italicised. Not significant results are shown in red.

**Extended Data Table 3.** Univariate cox regression Results of univariate cox models showing the relationship between implementations of the foundation model and the supervised methods and survival on LUNG1 and RADIO datasets. The median split on the training dataset (HarvardRT) is used, also shown in Fig S4 in the Kaplan-Meier curves.

|  | LUNG1 |  |  | RADIO |  |  |
| --- | --- | --- | --- | --- | --- | --- |
|  | beta | HR (95% CI for HR) | p.value | beta | HR (95% CI for HR) | p.value |
| Foundation model as feature extractor | -0.44 | 0.65 (0.52-0.81) | <0.005 | -0.84 | 0.43 (0.23-0.82) | 0.01 |
| Foundation model fine-tuned | -0.39 | 0.68 (0.5-0.92) | 0.01 | -0.32 | 0.72 (0.26-2.01) | 0.53 |
| Supervised (fine-tuned) | -0.24 | 0.79 (0.64-0.98) | 0.03 | -0.43 | 0.65 (0.37-1.15) | 0.14 |
| Supervised (random initialization) | -0.22 | 0.80 (0.65-1.00) | 0.05 | 0.20 | 1.22 (0.59-2.53) | 0.59 |

**Extended Data Table 4.**
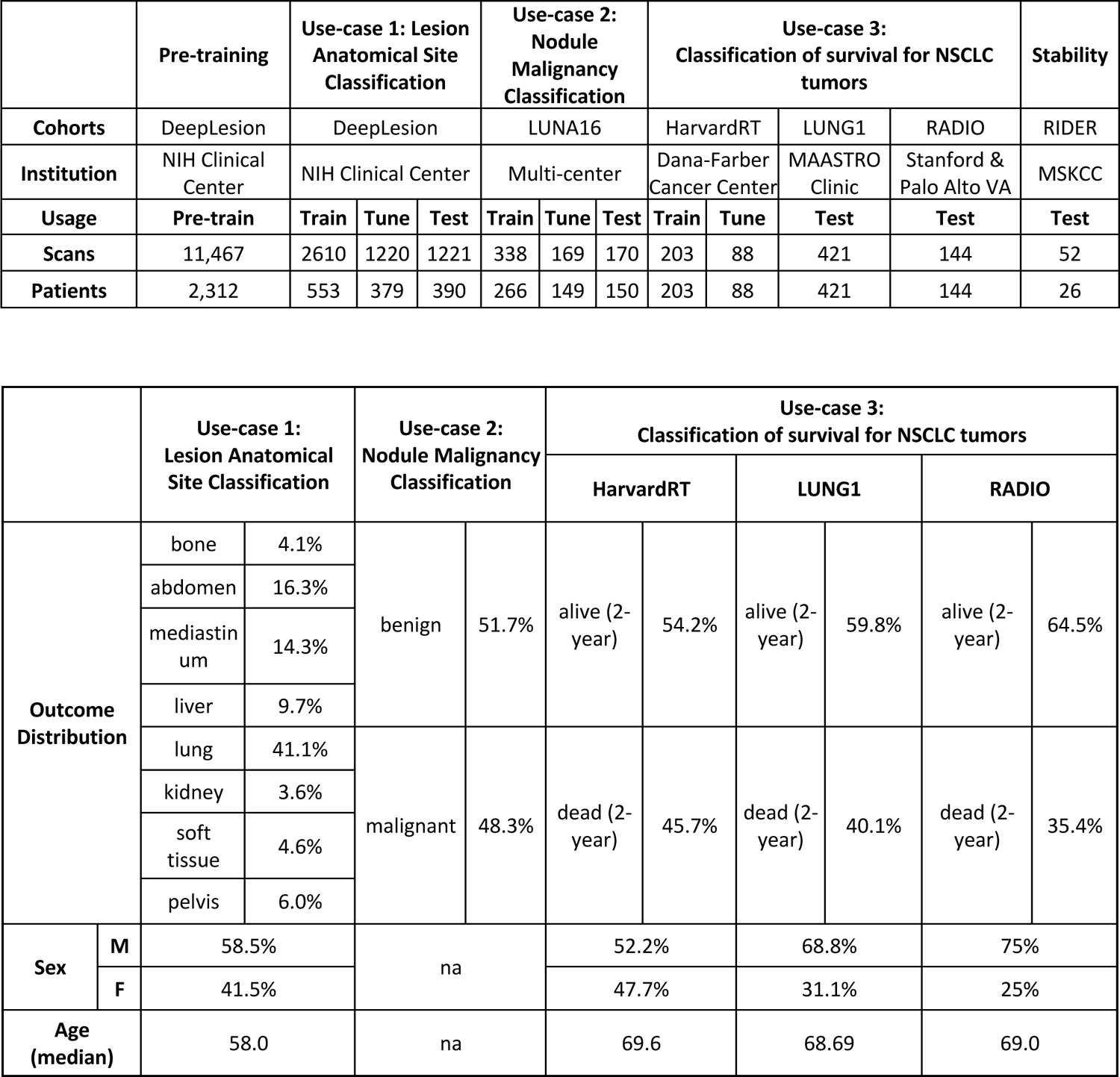
Dataset breakdown Table showing the 6 different cohorts used in this study along with eligible scans and patients used. A secondary table shows the outcome, sex, and age distribution of each of the cohorts.

## REFERENCES

1. Bommasani, R., et al. On the Opportunities and Risks of Foundation Models. *arXiv [cs.LG]* (2021).

2. Ouyang, L. et al. Training language models to follow instructions with human feedback. arXiv [cs.CL] 27730–27744 (2022).

3. Devlin, J., Chang, M.-W., Lee, K. & Toutanova, K. BERT: Pre-training of Deep Bidirectional Transformers for Language Understanding. arXiv [cs.CL] (2018).

4. Radford, A. et al. Learning transferable visual models from natural language supervision. *arXiv [cs.CV]* 8748–8763 (18--24 Jul 2021).

5. Chen, T., Kornblith, S., Norouzi, M. & Hinton, G. A Simple Framework for Contrastive Learning of Visual Representations. arXiv [cs.LG] (2020).

6. Oquab, M., et al. DINOv2: Learning robust visual features without supervision. *arXiv [cs.CV]* (2023).

7. Anja Thieme Microsoft Health Futures, United Kingdom et al. Foundation Models in Healthcare: Opportunities, Risks & Strategies Forward. doi:10.1145/3544549.3583177.

8. Moor, M. et al. Foundation models for generalist medical artificial intelligence. Nature 616, 259–265 (2023).

9. Mahajan, A. et al. Deep learning-based predictive imaging biomarker model for EGFR mutation status in non-small cell lung cancer from CT imaging. J. Clin. Orthod. 38, 3106–3106 (2020).

10. Hosny, A. et al. Deep learning for lung cancer prognostication: A retrospective multi-cohort radiomics study. PLoS Med. 15, e1002711 (2018).

11. Braghetto, A., Marturano, F., Paiusco, M., Baiesi, M. & Bettinelli, A. Radiomics and deep learning methods for the prediction of 2-year overall survival in LUNG1 dataset. Sci. Rep. 12, 14132 (2022).

12. Balestriero, R., et al. A Cookbook of Self-Supervised Learning. arXiv [cs.LG] (2023).

13. Huang, S.-C. et al. Self-supervised learning for medical image classification: a systematic review and implementation guidelines. NPJ Digit Med 6, 74 (2023).

14. Yan, K., Wang, X., Lu, L. & Summers, R. M. DeepLesion: automated mining of large-scale lesion annotations and universal lesion detection with deep learning. J Med Imaging (Bellingham) 5, 036501 (2018).

15. Zhao, B. et al. Evaluating variability in tumor measurements from same-day repeat CT scans of patients with non-small cell lung cancer. Radiology 252, 263–272 (2009).

16. Springenberg, J. T., Dosovitskiy, A., Brox, T. & Riedmiller, M. Striving for Simplicity: The All Convolutional Net. arXiv [cs.LG] (2014).

17. Smilkov, D., Thorat, N., Kim, B., Viégas, F. & Wattenberg, M. SmoothGrad: removing noise by adding noise. arXiv [cs.LG] (2017).

18. Hinshaw, D. C. & Shevde, L. A. The Tumor Microenvironment Innately Modulates Cancer Progression. Cancer Res. 79, 4557–4566 (2019).

19. Azizi, S., et al. Big Self-Supervised Models Advance Medical Image Classification. arXiv [eess.IV] (2021).

20. Krishnan, R., Rajpurkar, P. & Topol, E. J. Self-supervised learning in medicine and healthcare. Nat Biomed Eng 6, 1346–1352 (2022).

21. Ghesu, F. C. et al. Self-supervised Learning from 100 Million Medical Images. arXiv [cs.CV] (2022).

22. Haarburger, C., et al. Radiomics feature reproducibility under inter-rater variability in segmentations of CT images. Scientific Reports vol. 10 Preprint at 10.1038/s41598-020-69534-6 (2020).

23. Campello, V. M. et al. Minimising multi-centre radiomics variability through image normalisation: a pilot study. Sci. Rep. 12, 12532 (2022).

24. Kumar, D. et al. Discovery Radiomics for Pathologically-Proven Computed Tomography Lung Cancer Prediction. arXiv [cs.CV] (2015).

25. Lao, J. et al. A Deep Learning-Based Radiomics Model for Prediction of Survival in Glioblastoma Multiforme. Sci. Rep. 7, 10353 (2017).

26. Haarburger, C., Weitz, P., Rippel, O. & Merhof, D. Image-based Survival Analysis for Lung Cancer Patients using CNNs. arXiv [cs.CV] (2018).

27. Cho, H.-H. et al. Radiomics-guided deep neural networks stratify lung adenocarcinoma prognosis from CT scans. Commun Biol 4, 1286 (2021).

28. Taleb, A. et al. 3d self-supervised methods for medical imaging. Adv. Neural Inf. Process. Syst. 33, 18158–18172 (2020).

29. Tiu, E. et al. Expert-level detection of pathologies from unannotated chest X-ray images via self-supervised learning. Nat Biomed Eng 6, 1399–1406 (2022).

30. Zhou, Z. et al. Models Genesis: Generic Autodidactic Models for 3D Medical Image Analysis. Med. Image Comput. Comput. Assist. Interv. 11767, 384–393 (2019).

31. Chaitanya, K., Erdil, E., Karani, N. & Konukoglu, E. Contrastive learning of global and local features for medical image segmentation with limited annotations. arXiv [cs.CV] (2020).

32. Li, H. et al. Imbalance-Aware Self-supervised Learning for 3D Radiomic Representations. in Medical Image Computing and Computer Assisted Intervention – MICCAI 2021 36–46 (Springer International Publishing, 2021).

33. Li, Z., et al. A Novel Collaborative Self-Supervised Learning Method for Radiomic Data. arXiv [eess.IV] (2023).

34. Zhao, Z. & Yang, G. Unsupervised Contrastive Learning of Radiomics and Deep Features for Label-Efficient Tumor Classification. in Medical Image Computing and Computer Assisted Intervention – MICCAI 2021 252–261 (Springer International Publishing, 2021).

35. Parmar, C., Grossmann, P., Bussink, J., Lambin, P. & Aerts, H. J. W. L. Machine Learning methods for Quantitative Radiomic Biomarkers. Sci. Rep. 5, 13087 (2015).

36. Adebayo, J., Gilmer, J. & Muelly, M. Sanity checks for saliency maps. Adv. Neural Inf. Process. Syst. (2018).

37. Arun, N., et al. Assessing the Trustworthiness of Saliency Maps for Localizing Abnormalities in Medical Imaging. Radiol Artif Intell 3, e200267 (2021).

38. Setio, A. A. A. et al. Validation, comparison, and combination of algorithms for automatic detection of pulmonary nodules in computed tomography images: The LUNA16 challenge. Med. Image Anal. 42, 1– 13 (2017).

39. Kirby, J. NSCLC-Radiomics. https://wiki.cancerimagingarchive.net/display/Public/NSCLC-Radiomics.

40. Napel, S. NSCLC radiogenomics: Initial Stanford study of 26 cases. The Cancer Imaging Archive.

41. Wang, F. & Liu, H. Understanding the behaviour of contrastive loss. arXiv [cs.LG] 2495–2504 (2020).

42. Uemura, T., Näppi, J. J., Hironaka, T., Kim, H. & Yoshida, H. Comparative performance of 3D-DenseNet, 3D-ResNet, and 3D-VGG models in polyp detection for CT colonography. in Medical Imaging 2020: Computer-Aided Diagnosis vol. 11314 736–741 (SPIE, 2020).

43. Sohn, K. Improved deep metric learning with multi-class n-pair loss objective. Adv. Neural Inf. Process. Syst. 29, (2016).

44. Pedregosa, F. et al. Scikit-learn: Machine Learning in Python. arXiv [cs.LG] 2825–2830 (2012).

45. Akiba, T., Sano, S., Yanase, T., Ohta, T. & Koyama, M. Optuna: A Next-generation Hyperparameter Optimization Framework. in Proceedings of the 25th ACM SIGKDD International Conference on Knowledge Discovery & Data Mining 2623–2631 (Association for Computing Machinery, 2019).

46. Gmail, L. & Hinton, G. Visualizing Data using t-SNE. https://www.jmlr.org/papers/volume9/vandermaaten08a/vandermaaten08a.pdf?fbcl (2008).

47. Jorge Cardoso, M., et al. MONAI: An open-source framework for deep learning in healthcare. *arXiv [cs.LG]* (2022).

48. Gamer, M. irr: Various Coefficients of Interrater Reliability and Agreement. http://cran.r-project.org/web/packages/irr/irr.pdf (2010).

